# “If I am not aware then I don’t think the community is aware”: a mixed-methods study of health worker perspectives on future maternal group B streptococcus vaccination in Kenya

**DOI:** 10.1101/2025.06.07.25329190

**Authors:** Molly A. Sauer, Berhaun Fesshaye, Emily Miller, Jessica Schue, Prachi Singh, Rose Jalang’o, Nelly Bosire, Rosemary Njogu, Nicole Salazar-Austin, Ruth Karron, Rupali J. Limaye

**Affiliations:** Department of International Health, Johns Hopkins Bloomberg School of Public Health, Baltimore, Maryland, USA; National Vaccines and Immunization Program, Ministry of Health, Nairobi, Kenya; Thriving Teens Foundation, Nairobi, Kenya; Jhpiego, Nairobi, Kenya; Department of Pediatrics, Johns Hopkins School of Medicine, Baltimore, Maryland, USA; Department of Global and Community Health, George Mason University, Fairfax, Virginia, USA

**Keywords:** vaccination, pregnancy, group B streptococcus, acceptability, healthcare workers

## Abstract

Group B streptococcus (GBS) is a leading cause of neonatal and infant sepsis, pneumonia, and meningitis and is also associated with preterm birth, stillbirth, and other negative pregnancy outcomes. Infants born to mothers colonized with GBS bacteria are at greatest risk of the disease, and more than 95% of GBS disease occurs in sub-Saharan Africa. Screening and intrapartum antibiotic prophylaxis are important but insufficient prevention strategies, and their implementation is inconsistent or absent in many low- and middle-income settings. Maternal GBS vaccines are now in clinical trials, aiming to confer protection to newborns through maternal antibody transfer. Strengthening community uptake of these vaccines, once available, will hinge on strong delivery systems to reach pregnant persons, including providers who can provide appropriate and effective information and recommendations to their patients. We conducted a mixed-methods study to describe providers’ knowledge, attitudes, and beliefs related to GBS and maternal vaccination in Nakuru and Mombasa Counties, Kenya. Based on a cross-sectional survey of 100 providers and 12 in-depth interviews, we found that provider lack of awareness of GBS disease and its burden in the community is a substantial barrier to future GBS vaccine decision-making and demand; perceived community awareness of GBS was also limited. Actual and perceived provider knowledge of GBS varies, emphasizing the need for preemptive education and training in advance of a GBS vaccine introduction. Providers are trusted sources of information for their patients and are very likely to recommend a maternal vaccine that is recommended by the Ministry of Health. With licensed GBS vaccines on the horizon, there is an important opportunity to build provider awareness and knowledge of GBS risk, help strengthen provider and community trust in maternal vaccination and prevent serious GBS disease.

## Background

Group B streptococcus (GBS) is a bacterium (*Streptococcus agalactiae*) that is carried in gastrointestinal or urogenital tract of about one in three adults worldwide (1). While typically harmless, GBS can cause invasive disease in older adults, those with underlying medical conditions, pregnant people, and most significantly, newborns. Early onset GBS disease affecting newborns in the first week of life is especially serious and accounts for an estimated 60-70% of infant GBS disease (2,3).

GBS is a leading cause of neonatal and infant sepsis, pneumonia, and meningitis morbidity and mortality and is also associated with stillbirths and preterm births (2,4–8). Infants who survive may experience neurodevelopmental sequelae like speech, hearing, or vision issues; psychomotor impairment; and seizures (9). Data on GBS morbidity and mortality are limited, particularly in lower-resourced settings, but it is estimated to be associated with more than half a million preterm births (about 1 in every 27 preterm births), 46,000 stillbirths (about 1 in every 42 stillbirths), 91,000 newborn deaths (1 in every 25 newborn deaths), and 400,000 cases of early-onset (0-6 days of life) or late-onset (7-89 days) disease each year, in addition to lasting neurological implications among surviving infants (6,7,10–13).

An estimated one in five, or about 20 million, pregnant women carry GBS in the rectum or vagina; a systematic review and meta-analysis of 390 studies across 85 countries estimated that this rate varies regionally from 11-35% prevalence of rectovaginal GBS colonization among pregnant women (6–8). Newborns are at greatest risk of GBS disease if the mother is colonized with the bacterium; maternal infection is a prerequisite for early onset disease in the first 6 days of life. An estimated one-third of pregnant women carrying GBS transmit the infection to the newborn in utero or during delivery (10). GBS is most often asymptomatic in pregnant women but can still be transmitted to the developing fetus or newborn in the absence of maternal symptoms (6–8).

Low- and middle-income countries (LMICs) are home to more than 95% of GBS disease, with countries in sub-Saharan Africa facing the greatest burden, driven heavily by the absence of intrapartum antibiotic prophylaxis (IAP) policies (6,7,10,14). In Kenya, GBS morbidity and mortality data remain limited; two facility-based studies assessed prevalence of maternal colonization and GBS-associated negative birth outcomes with mixed results (15,16). A prospective cohort study by Seale, et al. (16) in urban, semi-rural, and rural areas of Mombasa and Kilifi Counties found about 12% prevalence of maternal colonization among the 7,967 women enrolled in the study, with colonization less common in women of lower socioeconomic status, and with high case fatality for early onset disease. The authors noted the likely underestimation of GBS burden, particularly at the community and outpatient care levels. A cross-sectional study by Jisuvei, et al. (15) described GBS colonization among 292 women seeking care in a large public referral hospital in Nairobi and found a 21% prevalence among participants, higher than the Seale study in coastal Kenya, but did not identify any risk factors associated with colonization. Both studies were facility-based; Seale and colleagues acknowledge that GBS burden in community and outpatient settings is likely underestimated due to culture insensitivity and limited case ascertainment due to rapid disease progression and lower access to care or facility delivery. This is an important area needing further study.

In many higher income settings, pregnant women are routinely screened for GBS in the third trimester and those diagnosed with or at high risk of GBS may be administered IAP— antibiotic treatment with penicillin or ampicillin—during labor and delivery to reduce the risk of neonatal infection and early onset disease (4,17). However, these measures will not prevent nosocomial or community-acquired late onset GBS disease and are insufficient for global control of GBS disease. Screening and IAP are costly and require adequate infrastructure and timely antenatal care (ANC) uptake, limiting their use in LMIC settings (4,8,14,17). A systematic review by LeDoare, et al. (18) found just three of 20 responding countries in sub-Saharan Africa reported an IAP policy and those that did have such a policy (including Kenya) indicated inconsistent and challenging implementation.

Given limited IAP use and treatment options, maternal vaccination has emerged as a high priority for prevention of GBS disease in newborns and was recently added to Gavi’s Vaccine Investment Strategy, signaling likely future co-financing support for introduction in LMICs (8,19–23). Maternal vaccination is the administration of a vaccine to a pregnant woman in order to confer passive immunity to the newborn via antibody transfer and/or to protect the mother from serious illness (24–28). Such vaccines are given to pregnant women, often in the second or third trimester, and antibodies developed in the vaccinated mother are passed transplacentally from mother to baby in utero; for infants who are breastfed, maternal antibodies can also be transferred via breastmilk (29). This passive immunity from the mother provides newborns with a crucial level of protection while their own immune systems are still developing and they remain at higher risk of serious disease. While the vaccines administered in pregnancy can vary by country and population, many national immunization programs include policies for tetanus toxoid-containing vaccines. Other routine maternal vaccines include influenza, pertussis, and COVID-19 vaccines, and most recently, vaccines for respiratory syncytial virus (RSV) were recommended for use in pregnancy by the World Health Organization (WHO) Strategic Advisory Group of Experts on Immunization (SAGE) (30).

Maternal GBS vaccine candidates aim to build immunity in pregnant women, allowing antibodies to be passed to the developing fetus so they are protected from GBS infection in utero, during delivery, and in the first few months of life (8,21,31–34). This is particularly important given the rapid progression of GBS disease in newborns and high case fatality rate in the first hours and days after birth, and for pregnant women and newborns in lower-resourced settings where screening, laboratory confirmation, and IAP are out of reach (31).

GBS vaccines are still several years away from availability (34). Currently, two candidates are in clinical trials (S1 Table). GBS6, a hexavalent conjugate vaccine by Pfizer, has entered Phase III trials and was granted “breakthrough status” from the U.S. Food and Drug Administration, allowing for accelerated review (32,35,36); and GBS-NN/NN2, an alpha-like protein fusion vaccine by Minervax, has completed Phase II trials (32,37–40). In anticipation of future GBS vaccines, Gavi, the Vaccine Alliance, has approved in principle the inclusion of GBS vaccines in its investment strategy—an important step to allow Gavi-eligible LMICs to begin considering if and how they will prioritize introduction of GBS vaccines in their immunization portfolios (23).

As maternal GBS vaccines are now on the horizon, there is an important window of opportunity to prepare LMIC decision-makers, health providers and systems, and communities for their introduction (41). In many LMIC settings, maternal vaccines face a range of challenges, including limited context-relevant evidence of disease burden and vaccine use, overburdened health systems, variable uptake of and access to antenatal care, limited delivery platforms for vaccination in pregnancy, competing national immunization program priorities, and hesitancy among pregnant women and their families (34,41–45).

Health care providers play a critical role in their patients’ decisions to accept vaccines in pregnancy, serving as arbiters of information, trusted partners in health decisions, sounding boards for concerns and questions, and sources of guidance (45–47). A strong, unequivocal, and consistent provider recommendation to be vaccinated with approved maternal vaccines— accompanied by information about the target disease and vaccine—is frequently cited as essential to maternal vaccine acceptance, across all antigens (20,45,48–54). To better equip health providers to answer patient questions and confidently make maternal vaccination recommendations, understanding the baseline knowledge of GBS disease and vaccines among providers and addressing key gaps in advance of vaccine availability may help strengthen willingness and confidence in recommendations once GBS vaccines are approved.

We sought to describe health care provider views on and experiences with GBS disease in pregnant women and newborns and their perceptions about GBS vaccination in pregnancy in preparation for maternal GBS vaccine availability in Kenya.

## Methods

### Study design and ethical approval

We conducted a mixed-methods study using a convergent parallel design. Quantitative data from a cross-sectional survey and qualitative data from in-depth interviews (IDIs) were collected concurrently from unique respondents. Survey and interview guide items were informed by various models, including the Health Belief Model and the WHO Behavioral and Social Determinants (BeSD) framework (55,56) and adapted for the study’s focus on maternal vaccination (S2 Table). All instruments were pre-tested and modified to ensure appropriate local terminology and phrasing, including use of the term “pregnant women”. While most people who can become pregnant are cisgender women who were born and identify as female, maternal vaccination topics are also relevant to the experiences and perspectives of transgender and other gender diverse people with the capacity to become pregnant. We have used the term “pregnant women” in this study and paper to reflect the language used by respondents and in consultation with local study partners on culturally appropriate terms. Data collectors were trained on survey and interview methods and best practices, ethical research practice, data management, and basic immunization and maternal health concepts, in addition to extensive practice administering each study instrument.

This work was conducted as part of the Maternal Immunization Readiness Initiative (MIRI), a multi-method study aiming to describe demand and readiness for maternal COVID-19, RSV, and GBS vaccines in Kenya. The MIRI study gathered insights from pregnant and lactating women, community members, health care providers, and policymakers; patient, community, and policy perspectives on maternal GBS vaccination are described separately (57–63).

The study received ethical approval from institutional review boards (IRB) at the Johns Hopkins Bloomberg School of Public Health (JHSPH), the Kenya Medical Research Institute (KEMRI), and the National Commission for Science, Technology & Innovation (NACOSTI). Data collection was conducted with the approval of the Kenya Ministry of Health and county health authorities in Nakuru and Mombasa counties. All participants provided oral informed consent.

### Study setting

The study was conducted in Nakuru and Mombasa Counties in Kenya. Nakuru County, located west of Nairobi in the Rift Valley, is home to Kenya’s fourth largest city and covers an area of nearly 7,500 km^2^, with more than half of the county’s population of 2.1 million classified as rural (64,65). Mombasa County, in the far southeast along the Indian Ocean coast, is a geographically small (220km^2^) but densely populated urban county of about 1.2 million (65,66).

We selected these two counties in consultation with study partners and Ministry of Health officials to maximize geographic and study population diversity. There is evidence that immunization coverage is often higher in urban areas than rural areas (67); we sought to explore how urban and rural providers’ perspectives may differ as well. According to Kenya’s most recent Demographic and Health Survey (65), full immunization coverage (national schedule) among children 12-23 months of age was consistent between urban and rural areas (55.7%, 55.0%) and between Mombasa and Nakuru Counties (70.4%, 69.2%).

In each county, we identified 10 health facilities—ranging from level 2 (dispensaries and community outpatient units/clinics) to level 5 (specialized referral hospitals) and including both public and private facilities—in consultation with local study partners and national and county health officials (Fig 1). Facilities were selected to maximize diversity of participants, positing that providers serving smaller community facilities may have different perspectives than those at referral hospitals, and to facilitate adequate sample size and reasonable accessibility for data collectors. All invited facilities were informed by local health officials prior to initial study team visits. Quantitative data were collected in all 20 participating facilities and qualitative data were collected in a sub-set of facilities.

**Fig 1.**
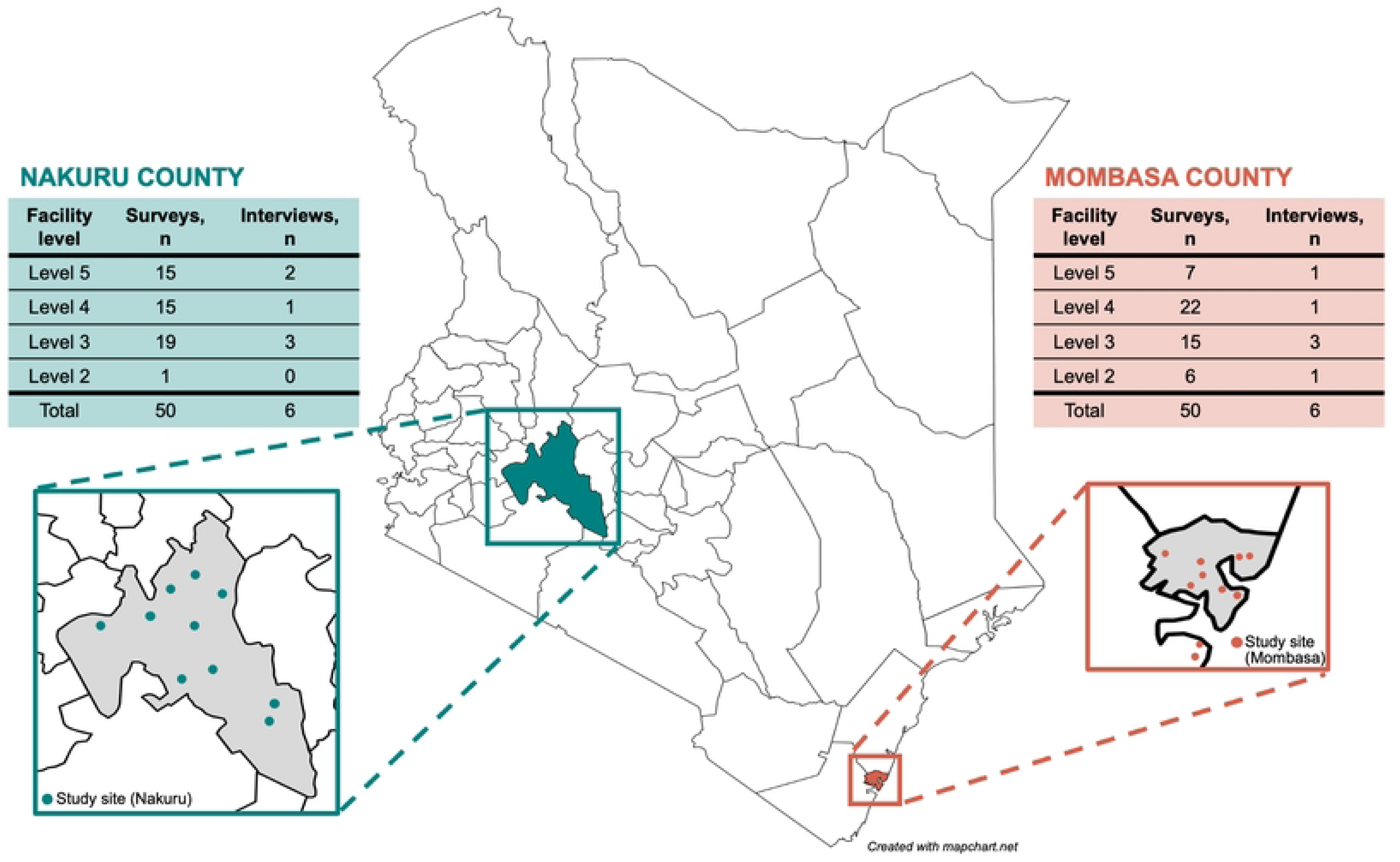
Study design and sampling frame.

### Data collection and analysis

Quantitative and qualitative data were collected concurrently from study facilities from July 25– September 21, 2022. Allocation of target number of surveys and interviews per facility within each county were determined based on facility catchment area and anticipated presence of providers involved in pregnancy-related care or immunization activities within the target data collection period. We used purposive sampling to enroll providers in each facility, prioritizing a range of perspectives and expertise relevant to future maternal GBS vaccine decision-making and demand. Providers were informed about the study and administered oral consent by trained data collectors; in accordance with IRB approvals, oral informed consent was administered in English in a private or semi-private area and was documented in survey responses or audio recordings. Survey and interview data were collected in English at or shortly after enrollment, depending on provider availability, and stored on encrypted servers using REDCap or OneDrive (39,40). Participants received a nominal reimbursement. Methods for the cross-sectional survey and IDIs are detailed below and summarized in Fig 2.

**Fig 2.**
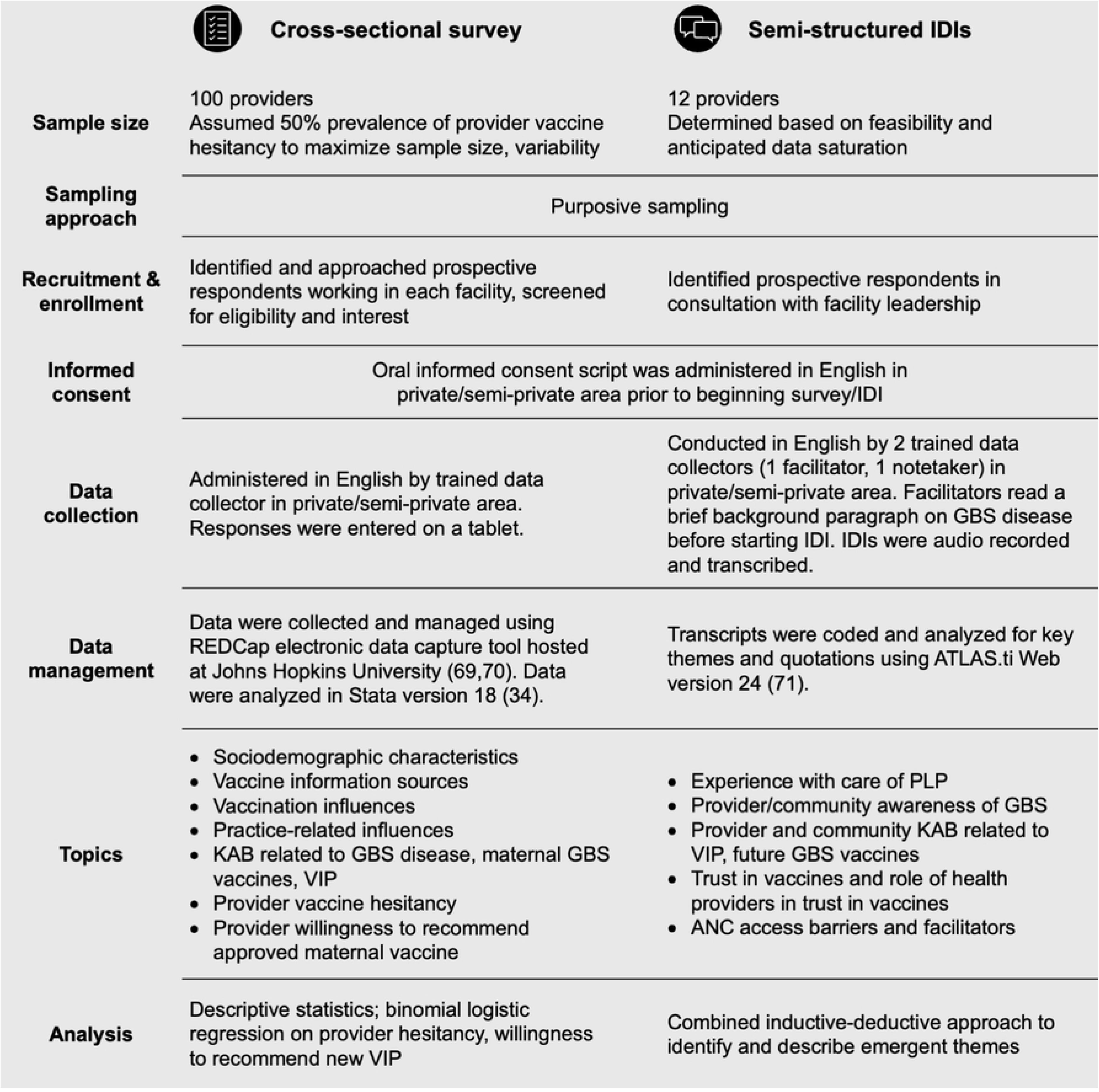
Summary of quantitative and qualitative methods.

### Quantitative methods

Cross-sectional surveys were administered by trained data collectors in REDCap using electronic tablets. Where appropriate, we adapted survey items from two validated vaccine hesitancy survey tools: the Parent Attitudes about Childhood Vaccines (PACV) questionnaire (68–70) and the WHO Vaccine Hesitancy Scale (VHS) (71). Survey items and response options are described in the supporting information (S2 Table).

Survey responses were analyzed to generate descriptive statistics to characterize respondents, stratifying by county as a proxy for rural/urban setting. We evaluated histograms of multi-level and Likert-like question responses to assess distribution; where appropriate, we collapsed or dichotomized categorical responses (i.e., “strongly agree or agree” and “strongly disagree or disagree”, “very strong or strong influence” and “very weak or weak influence”). We generated and dichotomized simple summative scores for relevant constructs with more than one item (i.e., perceived risk, supportive social norms, perceived safety, perceived benefits/vaccine effectiveness) and examined these variables at the construct and item levels.

To build on emergent themes identified through our mixed methods analysis, we conducted a simple logistic regression analysis to examine associations between sociodemographic factors, information sources, and vaccine-related knowledge, attitudes, and beliefs (KAB) with two outcomes of interest: provider vaccine hesitancy (2-item construct) and willingness to recommend a new, approved maternal vaccine (2-item construct). We generated a dichotomous summative score for vaccine hesitancy based on two items asked of all participants: “Thinking about your vaccine behavior when you were not pregnant, have you ever delayed getting a recommended vaccine or decided not to get a recommended vaccine for reasons other than illness or allergy?” and “Vaccines improve immunity. I believe it is better for me to develop my own immunity by getting sick rather than by getting a vaccine.”). Respondents who responded yes or don’t know (“Thinking about…”) or strongly agree/agree (“Vaccines improve…”) to one or both items were classified as “higher vaccine hesitancy” and those answering no or disagree/strongly disagree to both items were classified as “lower vaccine hesitancy.” To explore willingness (intentions) related to maternal vaccine recommendations, we dichotomized responses to two items: “Would you recommend an approved maternal vaccine if your head doctor recommended it?” (Very likely/likely, unlikely/very unlikely) and “Would you recommend an approved maternal vaccine if your head doctor did not recommend it?” (Very likely/likely, unlikely/very unlikely). We reviewed the resulting 2x2 table and generated a composite score for willingness to recommend in all cases. Respondents who answered “Very likely or likely” to both questions were classified as “higher willingness to recommend in all cases” and those answering “unlikely or very unlikely” to one or both questions were classified as “lower willingness to recommend in some/all cases”.

### Qualitative methods

We conducted semi-structured, in-person IDIs, which were audio recorded and transcribed. Following oral consent, facilitators read a brief background paragraph on GBS disease at the start of each IDI. Resulting qualitative data were analyzed for emergent themes using a combined inductive-deductive approach, aiming to identify unique themes from these data and align with relevant vaccine hesitancy and decision-making frameworks where appropriate. Team members not involved in facilitating interviews reviewed and independently coded one transcript, discussed proposed codes and themes, and generated a codebook; the codebook was applied to a second transcript, reviewed and refined, and then finalized to ensure agreement among team members and improve study rigor. The study team met regularly to discuss analysis and iteratively update the codebook. We double coded 15% of transcripts and estimated an inter-rater reliability of 93%.

## Results

We interviewed 12 providers and surveyed 100 providers. We used a convergent approach to analysis and interpretation of these data—quantitatively describing maternal vaccination knowledge, attitudes, and beliefs (KAB) among providers and more deeply examining potential associations using rich qualitative detail from the interviews. Here, we summarize respondent sociodemographic characteristics and then detail key themes emerging from our qualitative inquiry and descriptive analysis related to provider perspectives on maternal vaccination and GBS, with a focus on associations identified through the quantitative analysis and core components of relevant vaccine decision-making and vaccine hesitancy frameworks.

### Sociodemographic characteristics

Most health care providers surveyed (85%, n=85) identified as female, and 17 individuals reported themselves as pregnant (3% of all respondents) or lactating (14% of all respondents) (Table 1). A little more than half of respondents had at least one child and all had attained at least a college- or university-level education. Most respondents (70%) were enrolled at Level 3 or 4 facilities; 87% were enrolled at a public facility. Two-thirds (67%) of survey respondents were nurses, with an additional 21% identifying as a clinical officer. Other provider types included pediatricians, obstetrician/gynecologists, midwives, medical officers, nutritionists, laboratory technologists, or pharmacy technologists.

**Table 1.** Sociodemographic characteristics of quantitative sample, stratified by county (n=100)

| Factor | Total<br>(n=100) | Nakuru County<br>(rural)<br>(n=50, 50%) | Mombasa County<br>(urban)<br>(n=50, 50%) | *p* |
| --- | --- | --- | --- | --- |
| Gender |  |  |  |  |
| Female | 85 (85%) | 41 (82%) | 44 (88%) | 0.401 |
| Male | 15 (15%) | 9 (18%) | 6 (12%) |  |
| Age |  |  |  |  |
| 18-29 | 27 (27%) | 19 (38%) | 8 (16%) | 0.025 |
| 30-44 | 44 (44%) | 21 (42%) | 23 (46%) |  |
| 45+ | 29 (29%) | 10 (20%) | 19 (38%) |  |
| Pregnant/lactating status |  |  |  |  |
| Not pregnant or lactating | 68 (68%) | 33 (66%) | 35 (70%) | 0.803 |
| Pregnant | 3 (3%) | 1 (2%) | 2 (4%) |  |
| Lactating | 14 (14%) | 7 (14%) | 7 (14%) |  |
| N/A (male) | 15 (15%) | 9 (18%) | 6 (12%) |  |
| Number of children <18 years of age |  |  |  |  |
| None | 44 (44%) | 19 (38%) | 25 (50%) | 0.457 |
| One | 26 (26%) | 15 (30%) | 11 (22%) |  |
| Two or more | 30 (30%) | 16 (32%) | 14 (28%) |  |
| Education level |  |  |  |  |
| College/university completed | 93 (93%) | 49 (98%) | 44 (88%) | 0.050 |
| Post-graduate degree | 7 (7%) | 1 (2%) | 6 (12%) |  |
| Type/cadre of health worker |  |  |  |  |
| Nurse | 67 (67%) | 29 (58%) | 38 (76%) | 0.037 |
| Clinical officer | 21 (21%) | 11 (22%) | 10 (20%) |  |
| Doctor, midwife, medical officer | 5 (5%) | 3 (6%) | 2 (4%) |  |
| Other practitioner* | 7 (7%) | 7 (14%) | 0 (0%) |  |
| Facility type |  |  |  |  |
| Public | 87 (87%) | 43 (86%) | 44 (88%) | 0.766 |
| Private | 13 (13%) | 7 (14%) | 6 (12%) |  |
| Facility level |  |  |  |  |
| Level 5 | 23 (23%) | 16 (32%) | 7 (14%) | 0.042 |
| Level 4 | 36 (36%) | 15 (30 %) | 21 (42%) |  |
| Level 3 | 34 (34%) | 18 (36%) | 16 (32%) |  |
| Level 2 | 7 (7%) | 1 (2%) | 6 (12%) |  |

\*Other practitioners included nutritionists, pharmacy technologists, and laboratory technologists
Bold p values indicate statistical significance ( $p \leq 0.05$ )

Distribution of professional categories for interviewees was similar to the survey. More than half (n=7, 58%) of the 12 providers interviewed were nurses; we also interviewed three (25%) obstetricians/gynecologists and two (17%) clinical officers.

### Emergent themes related to maternal GBS vaccination

We identified several key themes from the qualitative data: knowledge and awareness of GBS, including its effects and prevention or treatment; perceived risk of GBS infection and severe outcomes; health promotion and trust in provider recommendations; maternal vaccination questions and concerns; provider hesitancy and information sources; and provider willingness to recommend new maternal vaccines. Generally, our qualitative findings aligned with survey findings but provided important nuance or context, including distinctions between provider knowledge and attitudes and their perceptions of community perspectives. Key quantitative and qualitative findings are presented in Fig 3 and detailed below; descriptive analysis of psychosocial factors among surveyed providers are presented in the supporting information (S3 Table).

**Fig 3.**
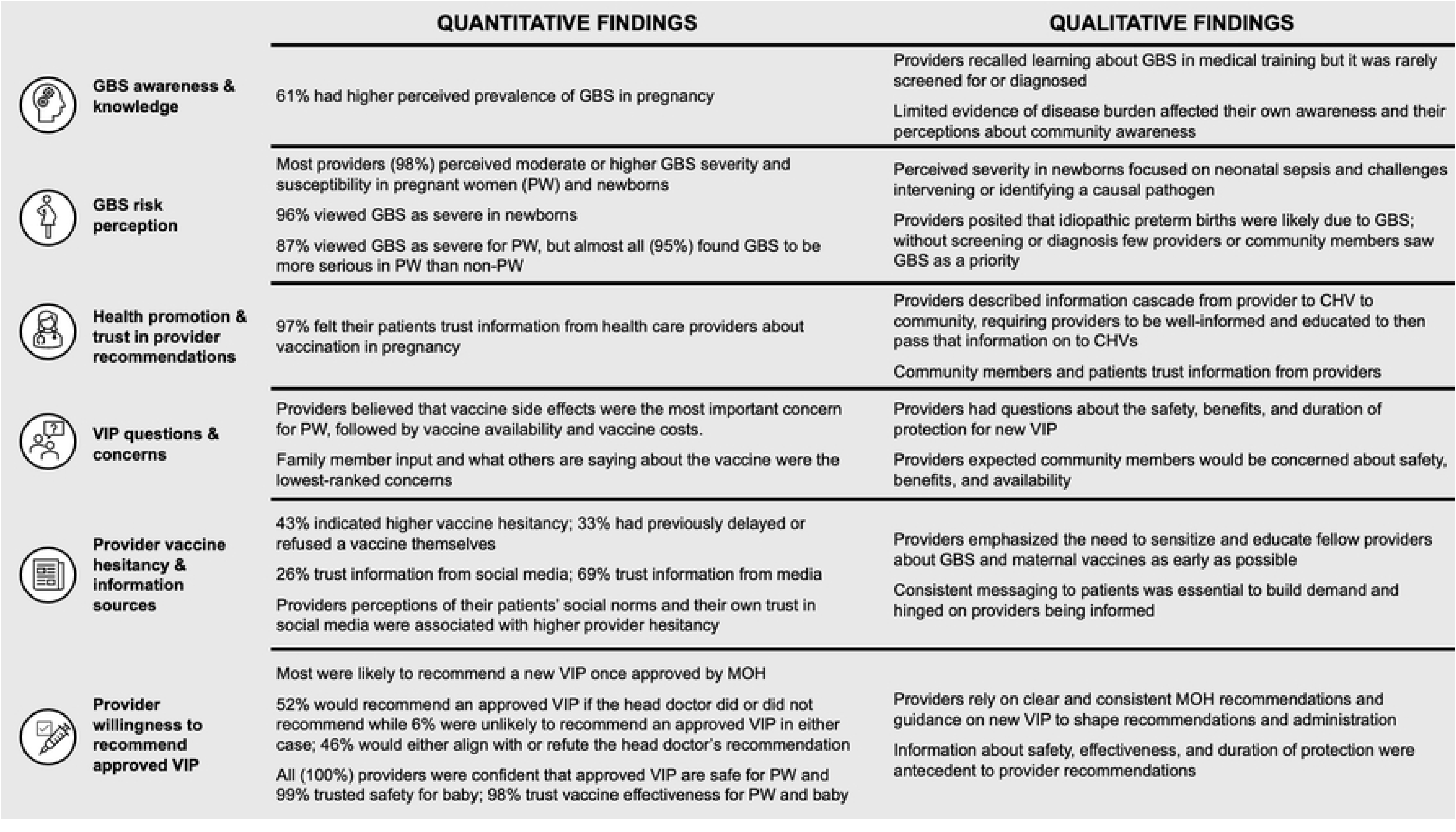
Joint display of quantitative and qualitative results by emergent theme.

#### GBS awareness and knowledge

Although nearly two-thirds (61%) of surveyed health care providers indicated a higher perceived prevalence of GBS infection among pregnant women (“The majority of pregnant women get GBS”), about one in three providers may either be unaware of GBS disease or may not believe that it affects most or many pregnant women. In qualitative interviews, again providers awareness and knowledge of GBS varied and an assumption was made that this limited knowledge extended to the communities they serve: *“If I am not aware then I don’t think the community is aware” (provider in Mombasa)*.

Respondents described the contributions of infections, including GBS, to neonatal morbidity and mortality but noted that that providers and community members are *“aware of the possibility of infection in general, maybe not specifically due to strep” (provider in Nakuru)*. Providers familiar with GBS had learned about the disease in their medical training but observed waning awareness, given other maternal health priorities:

> “*I heard of GBS when I was doing my medical training, but it was not emphasized though we read that in developed countries it is screened just before delivery. … the mother may not have symptoms and it does not have serious complications to the mother but to the pregnancy. The premature rupture of membranes, the premature labors and miscarriages, so because of that we don’t give it emphasis but close to delivery …. We learn it in medical school but when we come here we forget about it … In terms of the medical practitioners themselves, that awareness, that reminder that this GBS is not in their head. We concentrate on the five big killers.” (provider in Mombasa)*

Where providers were aware of GBS and its outcomes, they emphasized the challenge of timely diagnosis and intervention. They noted that GBS-specific screening and IAP are rare, and providers seldom have adequate information in time to intervene, only learning of GBS as a possible cause when examining a newborn with sepsis.

Respondents described a range of GBS-related syndromes and outcomes, as well as some less commonly associated with GBS (i.e., gastrointestinal illness, other pulmonary and cardiac conditions), noting that the disease is present albeit often unrecognized and undiagnosed. Providers highlighted this as a key barrier to GBS vaccine demand: *“The biggest problem here is the ignorance of the disease [among the community], because it is there and you know the impact of it is so overwhelming” (provider in Mombasa)*.

Taken together, our quantitative and qualitative findings demonstrate that GBS awareness is insufficient and highlight the need for more detailed insights to distinguish between real and perceived provider knowledge and awareness of GBS.

#### GBS risk perception

Nearly all survey participants (98%) indicated moderate or high perceived GBS risk, based on a three-item construct of perceived severity and susceptibility for mother and baby. There was overwhelming agreement about the risk of severe disease newborns but less consistency about the risk of GBS for pregnant women; 96% agreed or strongly agreed that GBS is dangerous for fetuses and babies, compared to 87% for pregnant women. Almost all respondents (95%) agreed or strongly agreed that GBS is more dangerous for pregnant women compared to non-pregnant women.

Despite high perceived severity and susceptibility among surveyed health care providers, qualitative results described more complex understandings of GBS risk. Several providers highlighted the challenge of appropriately understanding GBS severity (the degree to which GBS causes serious health outcomes in the mother or newborn) and susceptibility (the relative seriousness for GBS in pregnant vs. non-pregnant women). This was especially the case in settings where screening for GBS or a definitive diagnosis for causes of negative birth outcomes and maternal or neonatal infections are challenging and rare. This limited diagnostic or screening capacity for GBS was also highlighted as a barrier to communication between providers and with community members: *“Since we are not able to actually diagnose it, it is difficult to give it a name … we always suspect because … we don’t have a laboratory way of diagnosing it.” (provider in Nakuru)*.

Providers described high perceived prevalence and risk of GBS as related to preterm births and neonatal sepsis cases seen in health facilities:

> “*We have seen a lot of preterm births which we are not able to attribute to … a specific condition but we always suspect [GBS] because … it is widely known [as] a common one. So unless we are able to identify … all the preterm birth that come, we are able to test and actually say this is GBS, I suspect it may be a problem…they improve because we give [antibiotics] targeting … GBS. So when we see good change and improvement, looking backwards we can say maybe that was the GBS because the antibiotics were targeting the GBS.” (provider in Nakuru)*

Our survey results indicated providers’ high and consistent perceived risk of GBS in their pregnant and newborn patients, but qualitative findings described broader concern about infection in pregnancy and neonates, particularly sepsis. One provider emphasized that the absence of screening and diagnosis has limited providers’ awareness of the impact of GBS and its role in pregnancy complications and newborn morbidity and mortality, but that anything impacting those two populations would be prioritized:

> “*You see for the mother it is not a severe disease but for the pediatrics who are born with sepsis or pneumonia, maybe we should look at that burden. Maybe how many babies are born that could have gotten it from the mother. Maybe from that perspective, we would know the burden of GBS because for the mothers who are asymptomatic, maybe for babies who are born already with pneumonia and sepsis, but we don’t know what the cause is. …Yes of course [GBS] has an impact to the baby and to the pregnancy and to the outcome it has an impact to the community as well because we take maternal health very seriously for both mother and the newborn…so if it has severe complications to the newborn and the pregnancy outcomes, then of course we take it as important” (provider in Mombasa)*

#### Health promotion and trust in provider recommendations

Mirroring the findings from many studies on maternal vaccine acceptance and the importance of provider recommendations, 97% of surveyed respondents agreed or strongly agreed that their patients trust information from health workers about vaccination in pregnancy.

These findings were echoed and amplified in the qualitative results, as respondents consistently described the importance of health care providers in facilities and community health volunteers (CHVs) outside of facilities as trusted information and health promotion resources for patients and communities more broadly. Facility-level health providers were responsible for training, informing, and supporting CHVs and several described the importance of equipping CHVs to cascade this information to their communities. CHVs are often recognized as the backbone of Kenya’s primary health care system, providing education and information to community members and linking communities to health care services.

> “*We give [CHVs] info, we educate them, and we tell them now go to the community, spread this word that there is a new vaccine for … mothers or for babies. When they go in the villages, they are well accepted, because these are people that they know. They know them and they trust them. So normally when the – when [community members] are given info [from CHVs], they accept it.” (provider in Mombasa)*

In addition to CHVs, several providers described health talks or other health promotion sessions conducted in facilities for groups of pregnant women. These forums offered an opportunity to share information with community members and patients, answer questions, and build acceptance, as well as to hear from the community about rumors or concerns:

> “*If the health care worker is not informed, then he or she will give wrong information. Or he will say I even don’t know about that vaccine. Health care workers have to be equipped [and] use the CHVs to spread that word.” (provider in Mombasa)*

#### Maternal vaccine questions and concerns

Asked about questions related to future maternal GBS vaccines, interviewed providers consistently described their own need for information about side effects and safety, benefits, duration of protection, and operational aspects like storage and administration. Providers believed the community would mostly have questions about side effects, safety, benefits, and vaccine availability. These qualitative perspectives largely mirror how surveyed providers ranked seven concerns when asked which were most important (1) to least important (7) for their patients and communities (Fig 4). Providers consistently ranked side effects among the top concerns for pregnant or lactating women and communities, with an average ranking of 1.7 / 7, followed by availability, costs, and ingredients. Input from health workers, family, and others were seen as less critical, although 97% of providers surveyed indicated that their patients trust information from health providers about vaccines in pregnancy.

**Fig 4.**
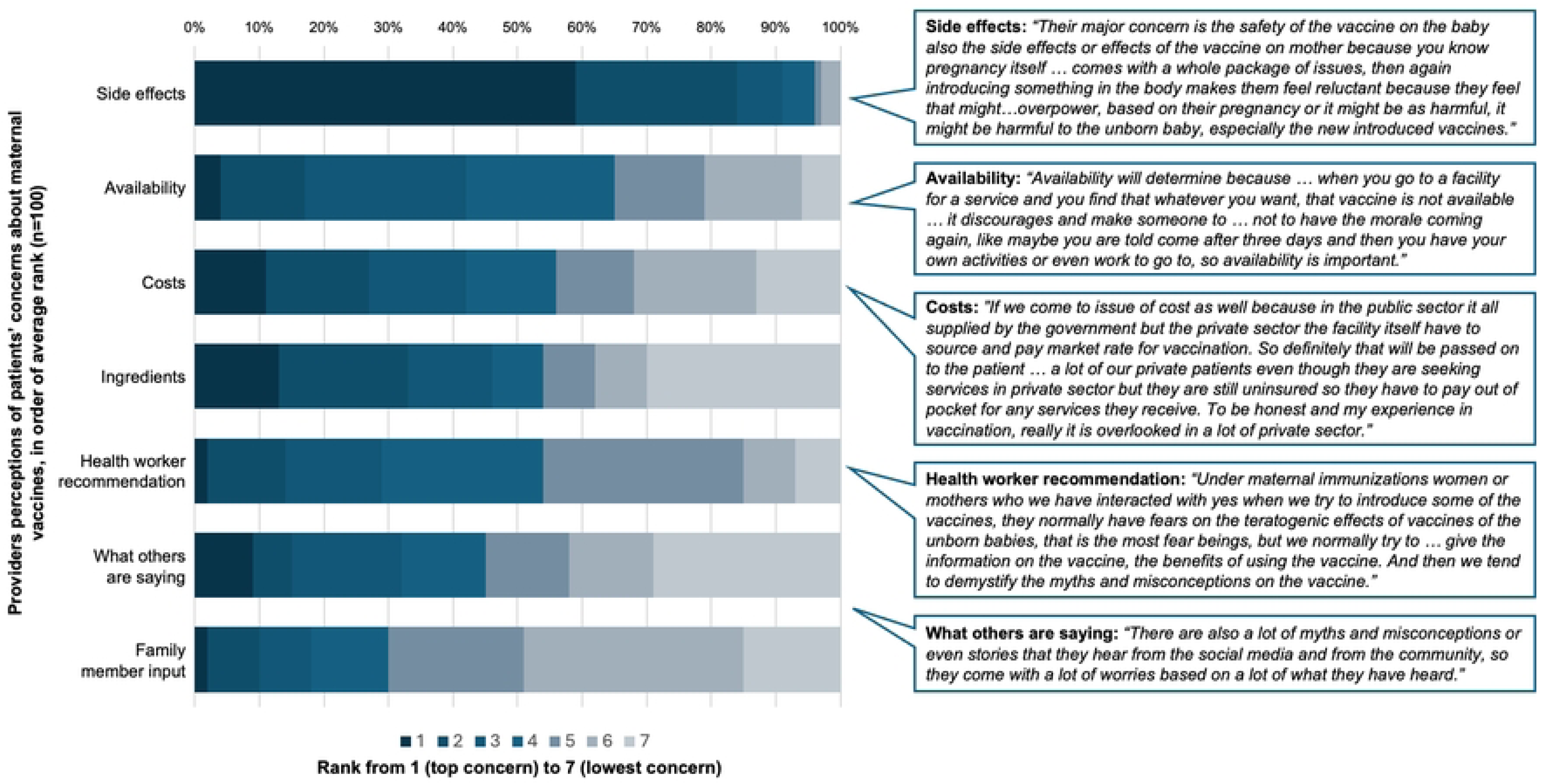
Provider ranking and illustrative quotations for patients’ top concerns about maternal vaccination concerns for their patients.

While input from the community or family members was generally not identified as a top concern, there was extensive discussion of community perspectives and the potential influence of rumors and misinformation on maternal GBS vaccine uptake among interview respondents,. Several interview respondents specifically discussed the influence of cultural and religious groups on vaccine acceptance. Survey respondents indicated relatively high supportive social norms (85%) for maternal GBS vaccination, based on the two-item construct. Asked to distinguish between what their communities believe people should do (injunctive norms) and what they would actually do (descriptive norms) when a maternal GBS vaccine is approved, slightly more providers agreed that their patients’ friends and families would encourage them to be vaccinated (93%) than agreed that patients’ friends and families would themselves be vaccinated with a maternal GBS vaccine (89%). Slightly fewer (86%) providers surveyed agreed or strongly agreed that their patients had some control over whether or not they get vaccinated during pregnancy (self-efficacy). This was statistically significantly higher among respondents in rural Nakuru County compared to urban Mombasa County (p = 0.021).

#### Provider vaccine hesitancy and information sources

Provider vaccine hesitancy was relatively high among surveyed providers—43% of providers had higher vaccine hesitancy based on a two-item construct assessing past delay or refusal of a vaccine and belief that illness-acquired immunity is better than vaccine-induced immunity. One in three (33%) had previously delayed or refused a vaccine (or did not know if they had). Three (18%) of the 17 pregnant or lactating health workers surveyed had delayed or refused a vaccine during pregnancy, and all three had also delayed or refused a vaccine when they were not pregnant; five (9%) of the 56 providers with children had previously delayed or refused a vaccine for their child, and all five had also previously delayed or refused a vaccine for themselves. Thirteen providers (13%) felt that developing immunity through illness was better than vaccine-induced immunity, another indicator of vaccine hesitancy.

Most providers relied on information from scientific or academic sources; 97% of surveyed providers trusted information they received from academics or scientists. Providers’ trust in media-derived information was lower, with 69% agreeing or strongly agreeing that they trust information received from media sources and 26% indicating they trust information received from social media.

Qualitative respondents did not describe delaying or refusing vaccination themselves. Many distinguished between strategies to educate and inform community members about new maternal vaccines and GBS disease and strategies to prepare, educate, and address reluctance among healthcare providers. The latter was a high priority for many respondents who discussed the importance of health education among the medical community to build provider awareness—recognizing the burden of GBS among pregnant women—and acceptability of a maternal vaccine as it nears availability. Providers were aligned in the need for proactive, consistent information and resources once a vaccine is on the horizon, with an emphasis on informing health workers so they are equipped to inform and educate their communities.

> “I think the earlier the better because this is a problem that we have…Health educate the health care worker, so that before the roll out of the vaccine, they can have enough info to be able to pass to the client, so that they can also accept the vaccine.” (provider in Nakuru)

> “*Human beings are free to do a lot of consultation…they even talk among themselves … or they may have friends who work somewhere else in the pharmacies, other clinics, so by the time they share the information with maybe us, the hospital has given to them before they accept, they must have had more information. They want to compare, do we [health care providers] point to one direction? If we [health care providers] are all going to the same direction they accept.” (provider in Nakuru)*

Considering the prevalence of hesitancy among providers alongside these qualitative results, we explored how provider KAB, information sources, and willingness to recommend future vaccines in pregnancy might relate to providers’ own history vaccine hesitancy (Table 2). We did not identify any significant associations between provider hesitancy and willingness to recommend an approved maternal vaccine. Providers who perceived less supportive descriptive social norms related to maternal vaccination within their communities were statistically significantly associated with higher vaccine hesitancy; the unadjusted odds of higher provider vaccine hesitancy were 4.1 (1.02, 16.57) times higher among providers who disagreed or strongly disagreed that the majority of their patient’s friends and family would get an approved maternal vaccine for GBS.

**Table 2.** Simple logistic regression analysis: factors associated with higher provider vaccine hesitancy (n=100)

| Factor | Associations with higher vaccine hesitancy<sup>a</sup> |  |  |
| --- | --- | --- | --- |
|  | OR | p-value | 95% CI |
| **Sociodemographic characteristics** |  |  |  |
| Gender |  |  |  |
| Female | Ref |  |  |
| Male | 0.86 | 0.80 | (0.28, 2.65) |
| Age |  |  |  |
| 18-29 | Ref |  |  |
| 30-44 | 0.90 | 0.83 | (0.34, 2.34) |
| 45+ | 0.57 | 0.30 | (0.19, 1.66) |
| County |  |  |  |
| Nakuru (rural) | Ref |  |  |
| Mombasa (urban) | 0.92 | 0.84 | (0.42, 2.03) |
| Number of children <18 years of age |  |  |  |
| None | Ref |  |  |
| One | 1.13 | 0.81 | (0.43, 2.99) |
| Two or more | 0.88 | 0.79 | (0.34, 2.25) |
| Highest education level attained |  |  |  |
| College/university completed | Ref |  |  |
| Post-graduate degree | 0.51 | 0.43 | (0.09, 2.75) |
| **Facility characteristics** |  |  |  |
| Facility level |  |  |  |
| Level 5 | Ref |  |  |
| Level 4 | 0.65 | 0.43 | (0.22, 1.91) |
| Level 3 | 1.65 | 0.36 | (0.57, 4.79) |
| Level 2 | 0.52 | 0.49 | (0.08, 3.26) |
| Facility ownership |  |  |  |
| Public | Ref |  |  |
| Private | 1.16 | 0.81 | (0.36, 3.73) |
| **Provider KAB** |  |  |  |
| The majority of pregnant women get GBS. [*prevalence*] |  |  |  |
| Strongly agree or agree | Ref |  |  |
| Disagree, strongly disagree, or don't know | 1.23 | 0.61 | (0.55, 2.78) |
| GBS is dangerous for pregnant women. [*severity*] |  |  |  |
| Strongly agree or agree | Ref |  |  |
| Disagree, strongly disagree, or don't know | 0.81 | 0.72 | (0.24, 2.66) |
| GBS is dangerous for fetuses and babies. [*severity*] |  |  |  |
| Strongly agree or agree | Ref |  |  |
| Disagree, strongly disagree, or don't know | 1.34 | 0.77 | (0.18, 9.92) |
| GBS is more dangerous for pregnant women compared to non-pregnant women. *[susceptibility]* |  |  |  |
| Strongly agree or agree | Ref |  |  |
| Disagree, strongly disagree, or don't know | 0.32 | 0.31 | (0.34, 2.93) |
| If there was an approved maternal vaccine for GBS, the majority of my patient's friends and family would get it. *[descriptive norm]* |  |  |  |
| Strongly agree or agree | Ref |  |  |
| Disagree or strongly disagree | 4.11 | 0.05 | (1.02, 16.57) |
| If there was an approved maternal vaccine for GBS, the majority of my patient's friends and family would encourage them to get it. *[injunctive norm]* |  |  |  |
| Strongly agree or agree | Ref |  |  |
| Disagree or strongly disagree | 1.85 | 0.44 | (0.39, 8.72) |
| My client has some control over whether or not they get vaccines during their pregnancy. *[self-efficacy]* |  |  |  |
| Strongly agree or agree | Ref |  |  |
| Disagree or strongly disagree | 0.31 | 0.09 | (0.08, 1.20) |
| If my clients need to visit a health facility for an appointment or a vaccine, my clients can easily go to that health facility. *[barriers]* |  |  |  |
| Strongly agree or agree | Ref |  |  |
| Disagree or strongly disagree | 1.34 | 0.77 | (0.18, 9.92) |
| I am confident that vaccines recommended for pregnant women during pregnancy are safe for the pregnant woman. |  |  |  |
| Strongly agree or agree | 1.00 |  | (1.00, 1.00) |
| I am confident that vaccines recommended for pregnant women during pregnancy are safe for the fetus and baby. |  |  |  |
| Strongly agree or agree | Ref |  |  |
| Disagree or strongly disagree | 1.00 |  | (1.00, 1.00) |
| If a new vaccine were approved for use among pregnant women, I trust that the vaccine would protect them. |  |  |  |
| Strongly agree or agree | Ref |  |  |
| Disagree or strongly disagree | 1.00 |  | (1.00, 1.00) |
| If a new vaccine were approved for use among pregnant women, I trust that the vaccine would protect the fetus and baby. |  |  |  |
| Strongly agree or agree | Ref |  |  |
| Disagree or strongly disagree | 1.00 |  | (1.00, 1.00) |
| Vaccine hesitancy (*construct, 2 items*) |  |  |  |
| Lower vaccine hesitancy | Ref |  |  |
| Higher vaccine hesitancy | 0.68 | 0.34 | (0.31, 1.51) |
| Information sources |  |  |  |
| My clients trust the information that they receive from their health care provider about vaccines during pregnancy. |  |  |  |
| Strongly agree or agree | Ref |  |  |
| Disagree or strongly disagree | 1.00 |  | (1.00, 1.00) |
| I trust the information provided by scientists and doctors at universities and academic institutions about vaccines during pregnancy. |  |  |  |
| Strongly agree or agree | Ref |  |  |
| Disagree or strongly disagree | 1.00 |  | (1.00, 1.00) |
| I trust the information that I have heard from the media (TV, radio, etc.) about vaccines during pregnancy. |  |  |  |
| Strongly agree or agree | Ref |  |  |
| Disagree or strongly disagree | 0.94 | 0.89 | (0.40, 2.21) |
| I trust the information that I have received from social media platforms (Facebook, WhatsApp) about vaccines during pregnancy. |  |  |  |
| Strongly agree or agree | Ref |  |  |
| Disagree or strongly disagree | 3.33 | 0.02 | (1.20, 9.24) |

<sup>a</sup> Health worker vaccine hesitancy was measured using two items asked of all respondents: "Thinking about your vaccine behavior when you were not pregnant, have you ever delayed getting a recommended vaccine or decided not to get a recommended vaccine for reasons other than illness or allergy?" and "Vaccines improve immunity. I believe it is better for me to develop my own immunity by getting sick rather than by getting a vaccine." Respondents who responded "Yes" or "Agree/strongly agree" to one or both items were classified as "higher vaccine hesitancy".

Lower reported trust in vaccine information from social media was also associated with provider vaccine hesitancy. The odds of higher vaccine hesitancy were 3.3 (1.20-9.24) times higher among providers reporting they did not trust the information they received from social media platforms (Facebook, WhatsApp) about vaccines during pregnancy. However, several providers still suggested engaging media and social media to disseminate messaging about new maternal vaccines before rumors can take hold and to facilitate demand, even as they acknowledged challenges with misinformation from these channels.

#### Provider willingness to recommend a new maternal vaccine

Almost all providers (97%) surveyed were likely or very likely to recommend a vaccine once approved by the Ministry of Health (MOH), but willingness to recommend an approved maternal vaccine varied based on recommendations from the clinic’s head doctor. When considering head doctor recommendation for or against the approved vaccine, just under half (48%) of providers were likely or strongly likely to recommend a new approved maternal vaccine regardless of whether the head doctor did or did not recommend the vaccine. We classified 52% of providers as reluctant (unlikely or very unlikely) to recommend a new approved maternal vaccine in some or all head doctor recommendation scenarios: 6% of respondents were unlikely to recommend an approved maternal vaccine regardless of whether the head doctor did or did not recommend it; 41% of respondents would adhere to head doctor recommendations for or against the vaccine; and 5% indicated they would counter or reject the head doctor recommendation (i.e., likely to recommend the vaccine when the head doctor did not, unlikely to recommend the vaccine when the head doctor did recommend it).

These findings generally aligned with our qualitative results, which emphasized the MOH policy in shaping when and how providers recommend or administer maternal vaccines. In general, most described adherence to national immunization policies and guidelines:

> “*It’s a national policy that they should be immunized. I don’t see anywhere whereby we can have an option that either they are supposed to be immunized or not… we give them the necessary health talk and then they accept to be given maybe for example… tetanus toxoid. So I don’t see where they are given the opportunity to say no.” (provider in Nakuru)*.

Some also described recommendations as part of their overall patient education—providing health information to patients rather than explicitly issuing a recommendation of their own. Providers acknowledged how clear policy guidance and provider-delivered health information encouraged acceptance of maternal vaccines.

> “*For pregnant patients, as long as the guidelines are clear and the information is given in details then I think I would recommend because at now the only vaccine that have for pregnant women is tetanus toxoid, that we give and I have not seen any client who is reluctant to take. So even if there is introduction of the others, as long as the information is given clearly then I think I will be in a position also to inform my clients about it.” (provider in Mombasa)*

Providers sought information on the safety profile and side effects, dosing schedule and duration of protection, and benefits to both mother and baby before they would be comfortable providing a recommendation and information to their patients:

> *“I’m the one who administers the vaccines, so if I have a negative opinion about the vaccine I would not encourage the clients to take the vaccine. If I have a positive attitude then that will give me strength to even convince or even explain further go ahead and give the impotence to the clients so that they can accept. I think I play a big role towards the vaccine administration and relaying the info to the clients.” (provider in Mombasa)*

While highlighting these key questions as antecedents for a recommendation, survey respondents overwhelmingly indicated that they trusted the safety and benefits (effectiveness) of approved vaccine in pregnancy for both mother and baby. When asked if they trusted that a new vaccine approved for use among pregnant women would protect the mother and, separately, the baby, 98% of respondents agreed or strongly agreed. Trust in safety was even higher: 100% agreed or strongly agreed that vaccines recommended for use in pregnancy were safe for the mother and 99% agreed or strongly agreed that they are safe for baby. Given high confidence in safety and effectiveness—the two key topics self-identified by qualitative respondents as driving recommendations—we investigated relationships between sociodemographic characteristics, KAB, and information sources and providers reluctance to recommend an approved maternal vaccine. The odds of being less willing to recommend an approved maternal vaccine in some or all cases were about three times higher (OR = 3.2, 1.07-9.70) among providers aged 45 or older, but no other variables were statistically significantly associated (p < 0.05) with being less willing to recommend an approved maternal vaccine (Table 3).

**Table 3.** Simple logistic regression analysis: factors associated with lower provider willingness to recommend an approved vaccine in pregnancy (VIP) (n=100)

| Factor | Associations with lower provider willingness to recommend VIP<sup>a</sup> |  |  |
| --- | --- | --- | --- |
|  | OR | p-value | 95% CI |
| **Sociodemographic characteristics** |  |  |  |
| Gender |  |  |  |
| Female | Ref |  |  |
| Male | 1.47 | 0.50 | (0.48, 4.48) |
| Age |  |  |  |
| 18-29 | Ref |  |  |
| 30-44 | 1.33 | 0.57 | (0.50, 3.50) |
| 45+ | 3.23 | 0.04 | (1.08, 9.70) |
| County |  |  |  |
| Nakuru (rural) | Ref |  |  |
| Mombasa (urban) | 0.85 | 0.69 | (0.39, 1.87) |
| Number of children <18 years of age |  |  |  |
| None | Ref |  |  |
| One | 1.28 | 0.62 | (0.48, 3.38) |
| Two or more | 1.43 | 0.45 | (0.56, 3.64) |
| Highest education level attained |  |  |  |
| College/university completed | Ref |  |  |
| Post-graduate degree | 6.13 | 0.10 | (0.71, 52.93) |
| **Facility characteristics** |  |  |  |
| Facility level |  |  |  |
| Level 5 | Ref |  |  |
| Level 4 | 0.48 | 0.18 | (0.16, 1.40) |
| Level 3 | 0.47 | 0.18 | (0.16, 1.41) |
| Level 2 | 0.71 | 0.70 | (0.13, 3.99) |
| Facility ownership |  |  |  |
| Public | Ref |  |  |
| Private | 2.30 | 0.19 | (0.66, 8.04) |
| **Provider KAB** |  |  |  |
| The majority of pregnant women get GBS. [*prevalence*] |  |  |  |
| Strongly agree or agree | Ref |  |  |
| Disagree, strongly disagree, or don't know | 0.68 | 0.35 | (0.30, 1.53) |
| GBS is dangerous for pregnant women. [*severity*] |  |  |  |
| Strongly agree or agree | Ref |  |  |
| Disagree, strongly disagree, or don't know | 1.09 | 0.89 | (0.34, 3.50) |
| GBS is dangerous for fetuses and babies. *[severity]* |  |  |  |
| Strongly agree or agree | Ref |  |  |
| Disagree, strongly disagree, or don't know | 0.29 | 0.30 | (0.03, 2.93) |
| GBS is more dangerous for pregnant women compared to non-pregnant women. *[susceptibility]* |  |  |  |
| Strongly agree or agree | Ref |  |  |
| Disagree, strongly disagree, or don't know | 0.22 | 0.18 | (0.02, 2.00) |
| If there was an approved maternal vaccine for GBS, the majority of my patient's friends and family would get it. *[descriptive norm]* |  |  |  |
| Strongly agree or agree | Ref |  |  |
| Disagree or strongly disagree | 0.74 | 0.65 | (0.21, 2.62) |
| If there was an approved maternal vaccine for GBS, the majority of my patient's friends and family would encourage them to get it. *[injunctive norm]* |  |  |  |
| Strongly agree or agree | Ref |  |  |
| Disagree or strongly disagree | 6.13 | 0.10 | (0.71, 52.93) |
| My client has some control over whether or not they get vaccines during their pregnancy. *[self-efficacy]* |  |  |  |
| Strongly agree or agree | Ref |  |  |
| Disagree or strongly disagree | 0.65 | 0.46 | (0.21, 2.04) |
| If my clients need to visit a health facility for an appointment or a vaccine, my clients can easily go to that health facility. *[barriers]* |  |  |  |
| Strongly agree or agree | Ref |  |  |
| Disagree or strongly disagree | 2.88 | 0.37 | (0.29, 28.65) |
| I am confident that vaccines recommended for pregnant women during pregnancy are safe for the pregnant woman. |  |  |  |
| Strongly agree or agree | 1.00 |  | (1.00, 1.00) |
| I am confident that vaccines recommended for pregnant women during pregnancy are safe for the fetus and baby. |  |  |  |
| Strongly agree or agree | Ref |  |  |
| Disagree or strongly disagree | 1.00 |  | (1.00, 1.00) |
| If a new vaccine were approved for use among pregnant women, I trust that the vaccine would protect them. |  |  |  |
| Strongly agree or agree | Ref |  |  |
| Disagree or strongly disagree | 1.00 |  | (1.00, 1.00) |
| If a new vaccine were approved for use among pregnant women, I trust that the vaccine would protect the fetus and baby. |  |  |  |
| Strongly agree or agree | Ref |  |  |
| Disagree or strongly disagree | 1.00 |  | (1.00, 1.00) |
| Vaccine hesitancy (*construct, 2 items*) |  |  |  |
| Lower vaccine hesitancy | Ref |  |  |
| Higher vaccine hesitancy | 0.68 | 0.34 | (0.31, 1.51) |
| Information sources |  |  |  |
| My clients trust the information that they receive from their health care provider about vaccines during pregnancy. |  |  |  |
| Strongly agree or agree | Ref |  |  |
| Disagree or strongly disagree | 1.88 | 0.61 | (0.16, 21.42) |
| I trust the information provided by scientists and doctors at universities and academic institutions about vaccines during pregnancy. |  |  |  |
| Strongly agree or agree | Ref |  |  |
| Disagree or strongly disagree | 0.45 | 0.52 | (0.04, 5.14) |
| I trust the information that I have heard from the media (TV, radio, etc.) about vaccines during pregnancy. |  |  |  |
| Strongly agree or agree | Ref |  |  |
| Disagree or strongly disagree | 1.18 | 0.70 | (0.50, 2.76) |
| I trust the information that I have received from social media platforms (Facebook, WhatsApp) about vaccines during pregnancy. |  |  |  |
| Strongly agree or agree | Ref |  |  |
| Disagree or strongly disagree | 1.11 | 0.81 | (0.46, 2.72) |

<sup>a</sup> Willingness to recommend a new VIP was measured using two items asked of all respondents: "Would you recommend an approved maternal vaccine if your head doctor recommended it?" and "Would you recommend an approved maternal vaccine if your head doctor did not recommend it?". Respondents who responded "Very likely or likely" to both questions were classified as "higher willingness to recommend in all cases" and those answering "unlikely or very unlikely" to one or both questions were classified as "lower willingness to recommend in some/all cases".

## Discussion

Our results suggest health care provider knowledge of GBS disease and maternal and neonatal infections is complex and variable. Surveyed providers reported high perceived GBS prevalence and risk, yet accompanying qualitative data illustrated how these perceptions are often shaped by other priority conditions and the limited antenatal screening and postnatal pathology to identify and address infections. Together, these data illustrate irregularities between providers’ actual awareness and knowledge of GBS disease and their perception or belief in their own awareness and knowledge, presenting a two-fold need and opportunity: first, to distinguish between real and perceived knowledge among providers in order to better shape educational and informational strategies, and second, to sensitize and train providers on GBS disease in advance of a vaccine to better equip them to communicate effectively with their patients and communities about both GBS disease and maternal vaccines. Discrepancies in descriptions of GBS disease and outcomes offer a potential target for provider training and education in advance of GBS vaccines, in order to cascade this knowledge and awareness to communities.

Few studies have examined provider perceptions of GBS disease and future maternal GBS vaccines and we are not aware of any other studies have taken a mixed methods approach, pairing survey data with in-depth interviews to allow for more nuanced interpretation. Qualitative inquiries on providers’ GBS vaccine perceptions have focused on high- or upper-middle-income settings (72,73). Many of our findings were similar to those identified in these studies—including variable knowledge among providers, strong emphasis on demonstrated safety, and the potential of a vaccine to mitigate limited diagnostic capacity—but generally our study saw more consistent support for a maternal GBS vaccine once recommended and available, with fewer caveats than these higher-resourced setting studies. Importantly, Patten (72) and Geoghegan (73) described how the varied provider attitudes toward future maternal GBS vaccines were linked to providers’ perceived value of vaccination vs. current screening and IAP strategies. GBS diagnostic testing and IAP are uncommon in Kenya and other LMICs, which may explain why our findings differ substantially from Patten and Geoghegan on this topic.

In LMIC settings, GBS disease and vaccines carry unique challenges. Screening and IAP are rare and inconsistent in these settings, requiring substantial health system infrastructure and resources to implement effectively. Without screening and laboratory confirmation, there is a dearth of local data and limited community and provider awareness of this disease and its impacts—prerequisites for effective vaccine rollout and uptake. New maternal RSV vaccines may help prime countries like Kenya for expanded maternal vaccination portfolios, but the unique complexities of GBS demand a targeted effort. While some providers indicated an awareness of GBS disease and its important implications in pregnancy and early infancy, others were unfamiliar or conflated GBS with other infections in childhood and older populations. Providers were consistent in the message that limited GBS diagnosis and screening in health facilities and communities and IAP unavailability heavily constrain their ability to confirm infection or intervene.

In general, providers were highly likely to recommend a new maternal GBS vaccine once approved by MOH and with adequate proof of its safety and effectiveness; providers own vaccine hesitancy appears unlikely to impact their willingness to recommend a new maternal vaccine. Providers’ trust in maternal vaccine safety and effectiveness is very high and most felt confident that, after an expected period of skepticism on introduction, most of their patients would be encouraged to accept a maternal GBS vaccine once approved. We observed that whether a facility’s head doctor recommends a new maternal vaccine once approved by the Ministry of Health appears to moderate providers’ recommendation intentions; more research is needed to understand these intra-facility dynamics and how they influence vaccine recommendations and acceptance. We also noted that providers 45 years of age or older were slightly less likely to recommend an approved maternal vaccine in all cases, compared to their younger colleagues. A study on non-maternal seasonal influenza vaccine use in China similarly found that older providers were less willing to recommend vaccination compared to younger colleagues (74). Further research is needed to understand why older providers appear to have reservations around recommending an MOH-approved maternal vaccine when factoring in head doctor recommendations for or against the vaccine; it will also be important to explore if these reservations extend to other non-maternal vaccines.

Providers in this study appeared to separate their personal and professional perspectives on vaccination, although more investigation into incongruent cases—where a provider has higher levels of vaccine hesitancy themselves but few or no reservations about recommending vaccination to their patients—could help shape education and health promotion strategies to build confidence and understand providers’ reasons for reluctance. Taken together, these qualitative and quantitative results emphasize the importance of clear and consistent maternal vaccination guidelines and policies from the MOH, including clear information on vaccine safety and benefits. The variability in how respondents would recommend or not recommend an approved maternal vaccine when considering head doctor recommendations demands further study, particularly to examine head doctor recommendations that conflict with MOH guidance.

Concerns and misperceptions were often tied to the exceptionalism of maternal vaccination. Stemming from the use of these vaccines only in pregnancy and why others are ineligible, exceptionalism has emerged as an important question among providers and other key stakeholders as new maternal RSV and potential GBS vaccines advance (61). Clearly and effectively explaining how maternal vaccines for infections acquired in utero or at (or shortly after) delivery work and why multiple different vaccines are needed for multiple diseases may be complicated. Providers noted that there will be questions among the community and beneficiaries, as well as providers, about why the vaccine is administered only during pregnancy and not to any other groups; these can contribute to rumors and myths. We detail questions from community members and pregnant and lactating women in a separate paper from this study (61) and our findings, described here and in Limaye et al. (61), generally align with other studies on maternal vaccine acceptability (44,52,54,75).

In particular, there is a persistent myth that vaccination in pregnancy affects fertility or is designed for population control; this rumor had particularly serious implications for maternal vaccination in Kenya and continues to circulate (76). Stakeholders in this study and others emphasize the need to continue to refute these myths while building acceptance for maternal immunization (61,62). Early communication about the risks of GBS disease and the safety and benefits of the new vaccine to community members and pregnant women will be critical to build trust, counter harmful misinformation, and clarifying how GBS vaccines differ from the more familiar tetanus toxoid-containing vaccines. We also noted higher perceived self-efficacy for maternal vaccination in Nakuru County compared to Mombasa County; this may be related to social, cultural, or religious differences between the two counties, but we did not gather data on these factors for each respondent. This could be an important area to explore further.

### Limitations and strengths

This study had several limitations. This study was conducted in Kenya, where there are ongoing efforts to prepare for future maternal vaccines. Our relatively small sample size likely contributed to large confidence intervals seen in the regression models and limited variance for several explanatory variables. Providers were enrolled at health facilities, thus excluding CHVs who are affiliated with but not based at these sites. This is an important area to explore further as Kenya prepares for new vaccines in pregnancy and seeks to reinforce the trust between communities and providers. Additionally, we strived to include a range of facilities in both urban and rural areas but were limited by logistical feasibility; it is reasonable to assume there are important differences related to maternal vaccine perspectives among providers in remote or harder-to-reach areas of Kenya. Our study adapted items from validated scales commonly used to understand parents’ perspectives on child vaccination in order to explore provider perspectives on maternal vaccination. Further research and validation are needed to determine if this adapted instrument can provide valid, reliable measurement of these concepts among providers and for maternal vaccines. While provider input is important in understanding and informing vaccine decisions and acceptance, provider perceptions of community perspectives may differ from actual community or patient perspectives. Social desirability bias is also likely.

As GBS vaccine candidates are still in clinical trials and several years from availability in public health programs, particularly in LMICs, we hypothesized that participants may be unfamiliar with GBS and provided a brief overview of GBS disease and vaccines at the start of each survey and interview. This may have influenced respondents to draw from the initial description rather than their own awareness or knowledge. We noted that GBS may be confused with other syndromes or conditions with similar names but different etiologies and interventions. In the context of this study, we posit that this is largely due to use of the term “streptococcus” or “strep,” which may be conflated with *Streptococcus pneumoniae* or other pathogens, and with the abbreviation “GBS” which was at times interpreted as Guillain-Barré Syndrome or other similarly abbreviated conditions. We acknowledge that this likely influenced survey and interview responses and our subsequent interpretation.

This study also has several strengths. It is among the first studies to explore provider perspectives on maternal vaccination in an LMIC setting, and to do so using a mixed-methods approach to marry quantitative measures of vaccine acceptance with rich context and detail of qualitative data. This allows for a more holistic and nuanced view of how providers think and feel about maternal vaccination in these spaces. Our variables were grounded in established social and behavioral frameworks for vaccine hesitancy and emphasized the most important drivers of acceptance. We included a diverse sample of providers and facilities, reflecting the range of practitioners involved in maternal and newborn care in LMIC settings.

## Conclusion

LMICs frequently introduce new vaccines years later than higher income settings (77) and are rarely prioritized for pre-licensure demand research like this study. While it will be several years before the earliest anticipated availability of maternal GBS vaccines, but this presents an opportunity to better understanding how upcoming maternal vaccines could be integrated into national immunization programs and how they might be perceived by those essential to their administration—health care providers, community health workers, and communities—to tailor training, communication, and other preparations for an eventual maternal GBS vaccine. Global, regional, and national scientific and clinical partners should work to strengthen health provider knowledge about GBS and upcoming vaccines and begin to build demand within the areas they serve. This preparation to build provider trust in maternal GBS vaccines—and, through them, community acceptance—is essential to ensure they remain trusted and informed resources on vaccination in pregnancy and can effectively prime patients, communities, and fellow providers for the eventual arrival of GBS vaccines in their facilities.

## Data Availability

Relevant quantitative data are within the paper and its supporting information. Qualitative data underlying the study are generated from semi-structured, in-depth interviews with health care providers in Kenya. Given the sensitivity of this information, and to protect participant confidentiality, data are not publicly available. In accordance with the informed consent obtained from participants at the time of data collection, interested parties can request de-identified transcripts or survey data by contacting the Johns Hopkins University Bloomberg School of Public Health Institutional Review Board all requests for access to the underlying study data will be reviewed and approved upon reasonable request.

## Acknowledgements

The authors are sincerely grateful for the health providers who willingly provided their time, expertise, and insights for this study, and to the administrators and health officials who facilitated this study’s conduct. The authors also acknowledge the contributions of the Jhpiego Kenya study management team and data collectors who expertly administered surveys and interviews to health providers and oversaw data collection activities across the two study counties.

## Supporting information

**S1 Table. GBS vaccine candidates undergoing human trials**

**S2 Table. Survey items and response options**

**S3 Table. Knowledge, attitudes, and beliefs related to vaccines in pregnancy (VIP) among health workers, stratified by county (n=100)**

## References

1. Group B Streptococcus infection causes an estimated 150,000 preventable stillbirths and infant deaths every year [Internet]. [cited 2024 Dec 8]. Available from: https://www.who.int/news/item/05-11-2017-group-b-streptococcus-infection-causes-an-estimated-150-000-preventable-stillbirths-and-infant-deaths-every-year

2. Le Doare K, Heath PT. An overview of global GBS epidemiology. Vaccine. 2013 Aug 28;31 Suppl 4:D7–12.

3. Edmond KM, Kortsalioudaki C, Scott S, Schrag SJ, Zaidi AK, Cousens S, et al. Group B streptococcal disease in infants aged younger than 3 months: systematic review and meta-analysis. The Lancet. 2012 Feb 11;379(9815):547–56.

4. Prevention of Group B Streptococcal Early-Onset Disease in Newborns: ACOG Committee Opinion, Number 797. Obstetrics & Gynecology. 2020 Feb;135(2):e51.

5. Alotaibi NM, Alroqi S, Alharbi A, Almutiri B, Alshehry M, Almutairi R, et al. Clinical Characteristics and Treatment Strategies for Group B Streptococcus (GBS) Infection in Pediatrics: A Systematic Review. Medicina. 2023 Jul;59(7):1279.

6. Russell NJ, Seale AC, O’Driscoll M, O’Sullivan C, Bianchi-Jassir F, Gonzalez-Guarin J, et al. Maternal Colonization With Group B Streptococcus and Serotype Distribution Worldwide: Systematic Review and Meta-analyses. Clin Infect Dis. 2017 Nov 15;65(Suppl 2):S100–11.

7. Gonçalves BP, Procter SR, Paul P, Chandna J, Lewin A, Seedat F, et al. Group B streptococcus infection during pregnancy and infancy: estimates of regional and global burden. Lancet Glob Health. 2022 Jun;10(6):e807–19.

8. World Health Organization. Group B Streptococcus (GBS). 2023 [cited 2023 Jan 11]. GBS vaccine pipeline. Available from: https://www.who.int/teams/immunization-vaccines-and-biologicals/diseases/group-b-streptococcus-(gbs)

9. Heymann DL, editor. Control of Communicable Diseases Manual. 20th edition. Washington, DC: American Public Health Association; 2015.

10. Seale AC, Bianchi-Jassir F, Russell NJ, Kohli-Lynch M, Tann CJ, Hall J, et al. Estimates of the Burden of Group B Streptococcal Disease Worldwide for Pregnant Women, Stillbirths, and Children. Clin Infect Dis. 2017 Nov 6;65(suppl_2):S200–19.

11. Preterm birth [Internet]. [cited 2024 Dec 8]. Available from: https://www.who.int/news-room/fact-sheets/detail/preterm-birth

12. Stillbirth [Internet]. [cited 2024 Dec 8]. Available from: https://www.who.int/health-topics/stillbirth

13. Newborn mortality [Internet]. [cited 2024 Dec 8]. Available from: https://www.who.int/news-room/fact-sheets/detail/newborn-mortality

14. Paul P, Gonçalves BP, Le Doare K, Lawn JE. 20 million pregnant women with group B streptococcus carriage: consequences, challenges, and opportunities for prevention. Curr Opin Pediatr. 2023 Apr;35(2):223–30.

15. Jisuvei SC, Osoti A, Njeri MA. Prevalence, antimicrobial susceptibility patterns, serotypes and risk factors for group B streptococcus rectovaginal isolates among pregnant women at Kenyatta National Hospital, Kenya; a cross-sectional study. BMC Infect Dis. 2020 Apr 22;20(1):302.

16. Seale AC, Koech AC, Sheppard AE, Barsosio HC, Langat J, Anyango E, et al. Maternal colonization with Streptococcus agalactiae and associated stillbirth and neonatal disease in coastal Kenya. Nat Microbiol. 2016 May 23;1(7):16067.

17. WHO recommendation on screening of pregnant women for intrapartum antibiotic prophylaxis for the prevention of early onset group B streptococcus disease in newborns [Internet]. Geneva: World Health Organization; 2024 [cited 2024 Dec 8]. Available from: https://www.who.int/publications/i/item/9789240099128

18. Le Doare K, O’Driscoll M, Turner K, Seedat F, Russell NJ, Seale AC, et al. Intrapartum Antibiotic Chemoprophylaxis Policies for the Prevention of Group B Streptococcal Disease Worldwide: Systematic Review. Clin Infect Dis. 2017 Nov 15 ;65(Suppl 2):S143–51.

19. World Health Organization. Group B streptococcus vaccine development technology roadmap: priority activities for development, testing, licensure and global availability of group B streptococcus vaccines [Internet]. 2017 [cited 2023 Jan 11]. Available from: https://www.who.int/publications-detail-redirect/WHO-IVB-17.10

20. Engmann C, Fleming JA, Khan S, Innis BL, Smith JM, Hombach J, et al. Closer and closer? Maternal immunization: current promise, future horizons. J Perinatol. 2020 Jun;40(6):844– 57.

21. World Health Organization. Urgent need for vaccine to prevent deadly Group B streptococcus [Internet]. 2021 [cited 2023 Jan 11]. Available from: https://www.who.int/news/item/02-11-2021-urgent-need-for-vaccine-to-prevent-deadly-group-b-streptococcus

22. Mantel C, Cherian T, Ko M, Malvolti S, Mason E, Giles M, et al. Stakeholder Perceptions About Group B Streptococcus Disease and Potential for Maternal Vaccination in Low- and Middle-Income Countries. Clinical Infectious Diseases. 2022 Jan 15;74(Supplement_1):S80–7.

23. Gavi. Vaccine Investment Strategy 2024 [Internet]. 2024 [cited 2024 Sep 3]. Available from: https://www.gavi.org/our-alliance/strategy/vaccine-investment-strategy-2024

24. Englund J, Paul Glezen W, Piedra PA. Maternal immunization against viral disease. Vaccine. 1998 Aug 1;16(14):1456–63.

25. Englund JA. Introduction. Maternal immunization - Promises and concerns. Vaccine. 2015 Nov 25;33(47):6372–3.

26. Chu HY, Englund JA. Maternal Immunization. Clinical Infectious Diseases. 2014 Aug 15;59(4):560–8.

27. Abbas AK, Lichtman AH, Pillai S. Basic immunology: Functions and disorders of the immune system. 4th ed. Philadelphia: Elsevier Saunders; 2014.

28. Kollmann TR, Marchant A, Way SS. Vaccination strategies to enhance immunity in neonates. Science. 2020 May 8;368(6491):612–5.

29. Etti M, Calvert A, Galiza E, Lim S, Khalil A, Le Doare K, et al. Maternal vaccination: a review of current evidence and recommendations. Am J Obstet Gynecol. 2022 Apr;226(4):459– 74.

30. World Health Organization. Meeting of the Strategic Advisory Group of Experts on Immunization, September 2024: conclusions and recommendations. Wkly Epidemiol Rec. 2024 Dec 6;99(49):719–40.

31. Seale AC, Baker CJ, Berkley JA, Madhi SA, Ordi J, Saha SK, et al. Vaccines for maternal immunization against Group B Streptococcus disease: WHO perspectives on case ascertainment and case definitions. Vaccine. 2019 Aug 14;37(35):4877–85.

32. Trotter CL, Alderson M, Dangor Z, Ip M, Le Doare K, Nakabembe E, et al. Vaccine value profile for Group B streptococcus. Vaccine. 2023 Nov 3;41:S41–52.

33. Absalon J, Simon R, Radley D, Giardina PC, Koury K, Jansen KU, et al. Advances towards licensure of a maternal vaccine for the prevention of invasive group B streptococcus disease in infants: a discussion of different approaches. Hum Vaccin Immunother. 2022 Dec 31;18(1):2037350.

34. World Health Organization, London School of Hygiene & Tropical Medicine. Group B Streptococcus Vaccine: full value vaccine assessment [Internet]. 2021 [cited 2023 Jan 11]. Available from: https://www.who.int/publications-detail-redirect/9789240037526

35. Madhi SA, Dangor Z, Heath PT, Schrag S, Izu A, Sobanjo-Ter Meulen A, et al. Considerations for a phase-III trial to evaluate a group B Streptococcus polysaccharide-protein conjugate vaccine in pregnant women for the prevention of early- and late-onset invasive disease in young-infants. Vaccine. 2013 Aug 28;31 Suppl 4:D52–57.

36. Pfizer. Trial To Evaluate The Safety, Tolerability, And Immunogenicity Of A Multivalent Group B Streptococcus Vaccine In Healthy Nonpregnant Women And Pregnant Women And Their Infants [Internet]. clinicaltrials.gov; 2024 Aug [cited 2024 Dec 10]. Report No.: NCT03765073. Available from: https://clinicaltrials.gov/study/NCT03765073

37. Minervax ApS. A Multi-centre Study to Evaluate the Safety, Tolerability and Immunogenicity of Two Doses of a Group B Streptococcus Vaccine (GBS-NN&#x2F;NN2) in Women Who Are Pregnant and Living With HIV and Women Who Are Pregnant and do Not Have HIV [Internet]. clinicaltrials.gov; 2023 Aug [cited 2024 Dec 10]. Report No.: NCT04596878. Available from: https://clinicaltrials.gov/study/NCT04596878

38. Minervax ApS. A Follow-up Trial to Assess the Persistence of the Immune Response to the Group B Streptococcus Vaccine (GBS-NN&#x2F;NN2) After a Primary Vaccination of Healthy Pregnant Women, and to Assess Safety, Reactogenicity, and Immunogenicity of the GBS-NN&#x2F;NN2 Vaccine When Administered During Follow-Up As a Booster Dose During a New Pregnancy [Internet]. clinicaltrials.gov; 2024 Sep [cited 2024 Dec 10]. Report No.: NCT06592586. Available from: https://clinicaltrials.gov/study/NCT06592586

39. Minervax ApS. A Multicentre, Multinational, Parallel Group, Observer-blind, Randomised, Placebo-controlled Study on the Group B Streptococcus Vaccine (GBS-NN&#x2F;NN2), Investigating the Immunogenicity and Safety of Four Vaccination Regimens in Pregnant Woman, Assessing IgG Specific to AlpN Proteins in Cord Blood and Maternal Blood, and the Safety Profile in Mother and Infant up to 6 Months Post-delivery [Internet]. clinicaltrials.gov; 2024 Mar [cited 2024 Dec 10]. Report No.: NCT05154578. Available from: https://clinicaltrials.gov/study/NCT05154578

40. Banks C, Lindbom BJ, Kitson G, Darsley M, Fischer PB. Preclinical development of a novel Group B (GBS) vaccine candidate for maternal immunization based upon the alpha-like protein family of GBS surface proteins (Alp). Birth Defects Research. 2023;115(9):933–44.

41. Sobanjo-ter Meulen A, Liljestrand J, Lawn JE, Hombach J, Smith J, Dickson KE, et al. Preparing to introduce new maternal immunizations in low- and lower-middle-income countries: A report from the Bill & Melinda Gates Foundation convening “Allies in Maternal and Newborn Care”; May 3–4, 2018. Vaccine. 2020 Jun 9;38(28):4355–61.

42. Geoghegan S, Shuster S, Butler KM, Feemster KA. Understanding Barriers and Facilitators to Maternal Immunization: A Systematic Narrative Synthesis of the Published Literature. Matern Child Health J. 2022;26(11):2198–209.

43. Davies B, Olivier J, Amponsah-Dacosta E. Health Systems Determinants of Delivery and Uptake of Maternal Vaccines in Low- and Middle-Income Countries: A Qualitative Systematic Review. Vaccines. 2023 Apr;11(4):869.

44. Otieno NA, Otiato F, Nyawanda B, Adero M, Wairimu WN, Ouma D, et al. Drivers and barriers of vaccine acceptance among pregnant women in Kenya. Hum Vaccin Immunother. 2020 Oct 2;16(10):2429–37.

45. MacDougall DM, Halperin SA. Improving rates of maternal immunization: Challenges and opportunities. Human Vaccines & Immunotherapeutics. 2016 Apr 2;12(4):857–65.

46. Dubé E. Addressing vaccine hesitancy: the crucial role of healthcare providers. Clinical Microbiology and Infection. 2017 May 1;23(5):279–80.

47. Krishnaswamy S, Lambach P, Giles ML. Key considerations for successful implementation of maternal immunization programs in low and middle income countries. Human Vaccines & Immunotherapeutics. 2019 Apr 3;15(4):942–50.

48. Collins J, Alona I, Tooher R, Marshall H. Increased awareness and health care provider endorsement is required to encourage pregnant women to be vaccinated. Hum Vaccin Immunother. 2014;10(10):2922–9.

49. Pathirana J, Nkambule J, Black S. Determinants of maternal immunization in developing countries. Vaccine. 2015 Jun 12;33(26):2971–7.

50. Kola-Palmer S, Keely A, Walsh J. “It has been the hardest decision of my life”: a mixed-methods study of pregnant women’s COVID-19 vaccination hesitancy. Psychol Health. 2023 May 22;1–21.

51. Mohammed H, Clarke M, Koehler A, Watson M, Marshall H. Factors associated with uptake of influenza and pertussis vaccines among pregnant women in South Australia. PLoS One. 2018;13(6):e0197867.

52. Nganga SW, Otieno NA, Adero M, Ouma D, Chaves SS, Verani JR, et al. Patient and provider perspectives on how trust influences maternal vaccine acceptance among pregnant women in Kenya. BMC Health Serv Res. 2019 Oct 24;19(1):747.

53. Karafillakis E, Francis MR, Paterson P, Larson HJ. Trust, emotions and risks: Pregnant women’s perceptions, confidence and decision-making practices around maternal vaccination in France. Vaccine. 2021 Jul 5;39(30):4117–25.

54. Lutz CS, Carr W, Cohn A, Rodriguez L. Understanding barriers and predictors of maternal immunization: Identifying gaps through an exploratory literature review. Vaccine. 2018 Nov 26;36(49):7445–55.

55. World Health Organization. Understanding the behavioural and social drivers of vaccine uptake, WHO position paper – May 2022. Wkly Epidemiol Rec. 2022 May 20;97(20):209– 24.

56. World Health Organization. Behavioural and social drivers of vaccination: tools and practical guidance for achieving high uptake [Internet]. World Health Organization; 2022. Available from: https://apps.who.int/iris/handle/10665/354459

57. Zavala E, Fesshaye B, Lee C, Mutwiwa S, Njagi W, Munyao P, et al. Lack of clear national policy guidance on COVID-19 vaccines influences behaviors in pregnant and lactating women in Kenya. Hum Vaccin Immunother. 2022 Oct 31;2127561.

58. Limaye RJ, Sauer M, Njogu R, Singh P, Fesshaye B, Karron RA. Characterizing Attitudes Toward Maternal RSV Vaccines Among Pregnant and Lactating Persons in Kenya: Key Considerations for Demand Generation Efforts for Vaccine Acceptance. Journal of the Pediatric Infectious Diseases Society [Internet]. 2023 Dec 16 [cited 2024 Oct 8];12(12). Available from: https://pubmed.ncbi.nlm.nih.gov/37944043/

59. Limaye RJ, Fesshaye B, Singh P, Karron RA. RSV awareness, risk perception, causes, and terms: Perspectives of pregnant and lactating women in Kenya to inform demand generation efforts for maternal RSV vaccines. Hum Vaccin Immunother. 2023 Aug;19(2):2258580.

60. Singh P, Fesshaye B, Lee C, Njogu RN, Karron RA, Limaye RJ. Maternal Immunization Decision-Making Among Pregnant and Lactating People in Kenya: A Qualitative Exploration of Peer Influences on Vaccine Decision-Making for a Future RSV Vaccine. Matern Child Health J. 2024 Oct;28(10):1822–32.

61. Limaye RJ, Singh P, Fesshaye B, Lee C, Schue J, Karron RA. “Why has this new vaccine come and for what reasons?” key antecedents and questions for acceptance of a future maternal GBS vaccine: Perspectives of pregnant women, lactating women, and community members in Kenya. Hum Vaccin Immunother. 20(1):2314826.

62. Limaye RJ, Fesshaye B, Singh P, Jalang’o R, Njogu RN, Miller E, et al. Understanding Kenyan policymakers’ perspectives about the introduction of new maternal vaccines. Health Policy and Planning. 2025 Jan 11;40(1):23–30.

63. Schue JL, Fesshaye B, Miller E, Singh P, Limaye RJ. COVID-19 vaccine preferences for pregnant and lactating women in Bangladesh and Kenya: a qualitative study. Front Public Health [Internet]. 2024 Aug 14 [cited 2025 Feb 3];12. Available from: https://www.frontiersin.org/journals/public-health/articles/10.3389/fpubh.2024.1412878/full

64. County Government of Nakuru. About County Government of Nakuru [Internet]. 2021 [cited 2024 Jul 28]. Available from: https://nakuru.go.ke/nakuru-county/

65. Kenya National Bureau of Statistics, ICF. Kenya Demographic and Health Survey 2022 [Internet]. Nairobi, Kenya, and Rockville, USA: KNBS and ICF; 2023 [cited 2024 Jul 28] p. 412. Available from: https://www.knbs.or.ke/wp-content/uploads/2023/07/Kenya-DHS-2022-Main-Report-Volume-2.pdf

66. Mombasa County Government. About Us – Mombasa County Government [Internet]. 2024 [cited 2024 Jul 28]. Available from: https://eservices.mombasa.go.ke/site/?page_id=64

67. Santos TM, Cata-Preta BO, Wendt A, Arroyave L, Blumenberg C, Mengistu T, et al. Exploring the “Urban Advantage” in Access to Immunization Services: A Comparison of Zero-Dose Prevalence Between Rural, and Poor and Non-poor Urban Households Across 97 Low- and Middle-Income Countries. J Urban Health. 2024 Jun 1;101(3):638–47.

68. Opel DJ, Taylor JA, Zhou C, Catz S, Myaing M, Mangione-Smith R. The Relationship Between Parent Attitudes About Childhood Vaccines Survey Scores and Future Child Immunization Status: A Validation Study. JAMA pediatrics. 2013 Nov;167(11):1065.

69. Opel DJ, Taylor JA, Mangione-Smith R, Solomon C, Zhao C, Catz S, et al. Validity and reliability of a survey to identify vaccine-hesitant parents. Vaccine. 2011 Sep 2;29(38):6598–605.

70. Opel DJ, Mangione-Smith R, Taylor JA, Korfiatis C, Wiese C, Catz S, et al. Development of a survey to identify vaccine-hesitant parents. Hum Vaccin. 2011 Apr;7(4):419–25.

71. Larson HJ, Schulz WS, Tucker JD, Smith DMD. Measuring vaccine confidence: introducing a global vaccine confidence index. PLoS Curr. 2015 Feb 25;7.

72. Patten S, Vollman AR, Manning SD, Mucenski M, Vidakovich J, Davies HD. Vaccination for Group B Streptococcus during pregnancy: Attitudes and concerns of women and health care providers. Social Science & Medicine. 2006 Jul 1;63(2):347–58.

73. Geoghegan S, Acosta F, Stephens LC, Gillan H, Valera S, Drew RJ, et al. Maternity care provider acceptance of a future Group B *Streptococcus* vaccine – A qualitative study in three countries. Vaccine. 2023 Mar 17;41(12):2013–21.

74. Ye L, Chen J, Fang T, Cui J, Li H, Ma R, et al. Determinants of healthcare workers’ willingness to recommend the seasonal influenza vaccine to diabetic patients: A cross-sectional survey in Ningbo, China. Human Vaccines & Immunotherapeutics. 2018 Dec 2;14(12):2979–86.

75. Otieno NA, Nyawanda B, Otiato F, Adero M, Wairimu WN, Atito R, et al. Knowledge and attitudes towards influenza and influenza vaccination among pregnant women in Kenya. Vaccine. 2020 Oct 7;38(43):6832–8.

76. Kenya Catholic Church tetanus vaccine fears “unfounded.” BBC News [Internet]. 2014 Oct 13 [cited 2024 Dec 17]; Available from: https://www.bbc.com/news/world-africa-29594091

77. Luthra K, Zimmermann Jin A, Vasudevan P, Kirk K, Marzetta C, Privor-Dumm L. Assessing vaccine introduction and uptake timelines in Gavi-supported countries: are introduction timelines accelerating across vaccine delivery platforms? BMJ Global Health. 2021 May 1;6(5):e005032.

